# Need for a definitive trial of local versus general anaesthesia in chronic subdural haematoma; lessons from a systematic review, survey, and scoping review of other surgical conditions

**DOI:** 10.1101/2025.07.20.25331843

**Authors:** Daniel J. Stubbs, Conor S. Gillespie, Matthew L. Watson, Basil Nourallah, Caroline M Phillips, George Gathercole, Jamie Brannigan, Keng Siang Lee, Orla Mantle, Vian Omar, Adele Mazzoleni, Githmi Palahepitiya Gamage, Alvaro Yanez Touzet, Munashe Veremu, Youssef Chedid, William H Cook, Karanjit Loyal, Gideon Adegboyega, Oliver D Mowforth, Edward Goacher, Apoorva Singh, Jonathan P. Coles, Alexis Joannides, Angelos Kolias, Judith Dinsmore, Iain Moppett, Michael Nathanson, Sally R Wilson, Amit Deshmukh, Edoardo Viaroli, David K. Menon, Ellie Edlmann, Benjamin M. Davies, Peter J Hutchinson, Improving Care In Elderly Neurosurgery Initiative (ICENI) Working Group

## Abstract

**Background:** Chronic subdural haematoma (cSDH) is a common neurosurgical condition, many patients have significant comorbidity or are living with frailty. Surgery is effective and can be performed under local anaesthesia (with or without sedation) or general anaesthesia. Optimal technique for both GA and LA is poorly defined but similar questions have been explored in other surgical settings. We sought to clarify the breadth of evidence for anaesthetic technique in cSDH surgery, while drawing on relevant literature from other disciplines to understand how a definitive trial of this question could be performed.

**Materials and Methods:** We used a combination of systematic and narrative literature search, review of trial registries, the Cochrane database, and a survey of anaesthetic and neurosurgical practitioners. An updated systematic review and meta-analysis of trial and observational studies in this area was performed following PROSPERO registration.

**Results:** We identified a paucity of high-quality studies, especially randomised trials, exploring this question. The literature, and a survey of anaesthetists and surgeons, suggest that local anaesthesia may bring benefits in shorter hospital stay and reduced complications. Registered studies in this field are single centre in nature while a synthesis of Cochrane reviews in other fields echoes issues of equipoise, study design, and outcome choice as key challenges in designing a definitive trial.

**Conclusions:** There is significant interest in this topic as evidenced by published and emerging literature and views of anaesthetists and surgeons. No registered trial is multi-centre or draws on challenges identified in similar trials from other disciplines. Our paper helps create a roadmap to a definitive trial of this crucial question.

## Introduction

A chronic subdural haematoma (cSDH) is a collection of altered blood beneath the dural membrane [1, 2]. It is associated with increasing age [3] and patients commonly have multiple co-morbidities or are living with frailty. Patients can present with a wide range of symptoms [4, 5]. cSDH may form with or without antecedent trauma [6]. Case numbers are projected to rise, with data from the United Kingdom (UK) and United States (US) suggesting the number of patient requiring operative management may rise by over 50% in the coming decades [2, 6, 7].

Treatment for symptomatic or large cSDH is primarily surgical [8, 9], although other methods have been investigated as alternative or adjuvant strategies [3, 10]. Although surgery is effective, perioperative morbidity including infection and recurrence can be significant [11]. Various surgical techniques are used, including cSDH evacuation via burr hole, mini-craniotomy, or twist drill craniostomy [12]. Surgery can be performed under local or general anaesthesia however evidence as to optimum technique is lacking [13–15]. This includes middle meningeal artery embolisation, with trial results yielding mixed results [16–18]. Previous systematic reviews have sought to answer this question [19, 20] although data are scarce. Consequently, practice varies, often by country, with UK practice highly skewed towards the use of general anaesthesia (93% of cases based on a 2016 national audit) [4].

In other disciplines, high-quality randomised controlled trials have sought to evaluate whether anaesthetic modality influences patient outcome [19]. Interpretation and conduct of such trials is difficult [21, 22] with challenges around patient selection, expertise, and equipoise. Although previous systematic reviews have revealed a paucity of high-quality randomised data examining anaesthetic modality for cSDH evacuation [20], a recent study has shown local anaesthetic use led to reduced complications and shorter hospital stay data [23]. Even in the context of equivalent longer-term outcomes, these short-term dividends may be meaningful for patients while offering productivity gains for pressured services [24]. Overall, this suggests a well-planned trial to answer this question is pressing.

Underpinned by a systematic literature search, joint surgical and anaesthetic practitioner survey, and a structured review of literature from other surgical disciplines, this article summarises current practice and explores why and how a comprehensive trial of anaesthetic modality for cSDH surgery might be conducted.

## Methods

### Overview

This paper summarises literature from both cSDH and other surgical disciplines to examine the need for, a randomised controlled trial (RCT) of local versus general anaesthesia for cSDH surgery. Evidence is presented from four sources: a systematic review and meta-analysis, a review of upcoming trials, a structured review of studies from other surgical disciplines, and a bespoke survey of surgical and anaesthetic practitioners.

### Systematic review of LA v GA in cSDH surgery

To inform new clinical practice guidelines for cSDH [15, 25], we conducted a systematic review of anaesthetic modality and outcome. Full methods and detailed results (such as risk of bias assessments and PROSPERO registration are available in the supplemental materials and other publications). Our findings are consistent with the findings of previously published systematic reviews on this topic [8, 19, 20]. We present the summary findings of the meta-analysis in Figure 1 for ease of reference, while full methods and supporting results are available in the supplemental material. The search built on previously published studies by describing the range of local anaesthesia and sedation (LAS) techniques used and synthesizing any head-to-head comparisons.

### Survey of anaesthetic and surgical practitioners

Paired surveys to examine the perspectives of both surgeons and anaesthetists on the use of LAS in cSDH surgery were created. Questions examined the frequency with which a respondent was exposed to LAS use, relevant monitoring or pharmacological techniques employed, factors perceived as favouring general (GA) or local anaesthesia, and views on the need for a definitive trial. Exact questions posed to surgeons and anaesthetists are provided in supplemental methods. Surveys were distributed by email to members of the Society for British Neurological Surgeons (SBNS) and Neuroanaesthesia and Critical Care Society (NACCS) between December 2021 and January 2022 after review by the councils of both organisations. Surveys were distributed using the SurveyGalaxy software (www.surveygalaxy.com). As this was an anonymised survey of healthcare professionals, ethical review was not required [26]. Quantitative results are summarised in the text and supplemental results. Free-text responses to each question were thematically coded and mapped as to whether they represented advantages or disadvantages of conducting surgery under LAS.

### Identification of relevant ongoing trials

We searched the WHO International Clinical Trials Registry Platform (ICTRP) [27], clinicaltrials.gov [28], and ISRCTN database to identify ongoing or registered trials [29]. Trial databases were searched for ‘Chronic Subdural’ and all entries reviewed. Studies were included if they explored elements of anaesthetic technique of relevance to the conduct of cSDH surgery and were included if their status was listed as ‘recruiting’, ‘unknown’, or ‘completed with no results’.

### Learning from other surgical conditions

We searched the Cochrane database on 15th January 2025 to identify reviews of trials comparing local or regional anaesthetic techniques to general anaesthesia in other surgical conditions. We used the search terms ‘local versus general anaesthesia’ OR ‘regional versus general anaesthesia’ and included any review that compared these modalities in any surgical or procedural discipline. We included both local and regional (e.g. spinal or peripheral nerve block) anaesthesia as comparisons as it was felt that both comparators may share core issues of relevance to trial design in this setting.

## Summary of examined evidence

### Systematic review and meta-analysis

Following our systematic search and screening, 15 studies were included to address the primary aim question (comparing LAS vs GA) while four additional studies were identified for the secondary aim of the systematic search – describing the range of LAS techniques and any head-to-head comparisons. Full details, PRISMA flow-chart and risk of bias assessments for each study are available in supplemental results. Summary results of meta-analyses against relevant outcomes shown in Figure1.

### Relationships between anaesthetic modality and outcomes in cSDH

The relationship between anaesthetic modality and outcomes including post-operative complications, recurrence, length of stay (LOS), and mortality are summarised in **Figure 1**. Broadly, our findings are comparable with other systematic reviews published on this topic [19, 20]. When restricted to studies with low risks of bias there appear to be a significant association between LAS and fewer postoperative complications (Odds ratio = 0.42 [95% CI 0.30, 0.59], P = 0.0008]) **(Figure 1)**. However, this was not seen when only RCTs (*n = 129 patients* total) were analysed (Odds ratio = 0.38 [95% CI 0, 1739], P = 0.39]). In a meta-analysis of six studies looking at length of hospital stay outcome, with a significant association between LAS and a shorter LOS (Mean difference = −2.49 [95%/ CI −3.66, −1.32] days, P < 0.0001]) although heterogeneity was significant (I^2^ = 87%). This is consistent with two studies demonstrating a shorter duration of anaesthesia and surgery (Mean difference = −29.52 [-37.27, −21.76] minutes P < 0.001]). No effect of LAS on recurrence of cSDH or mortality was seen.

**Figure 1.**
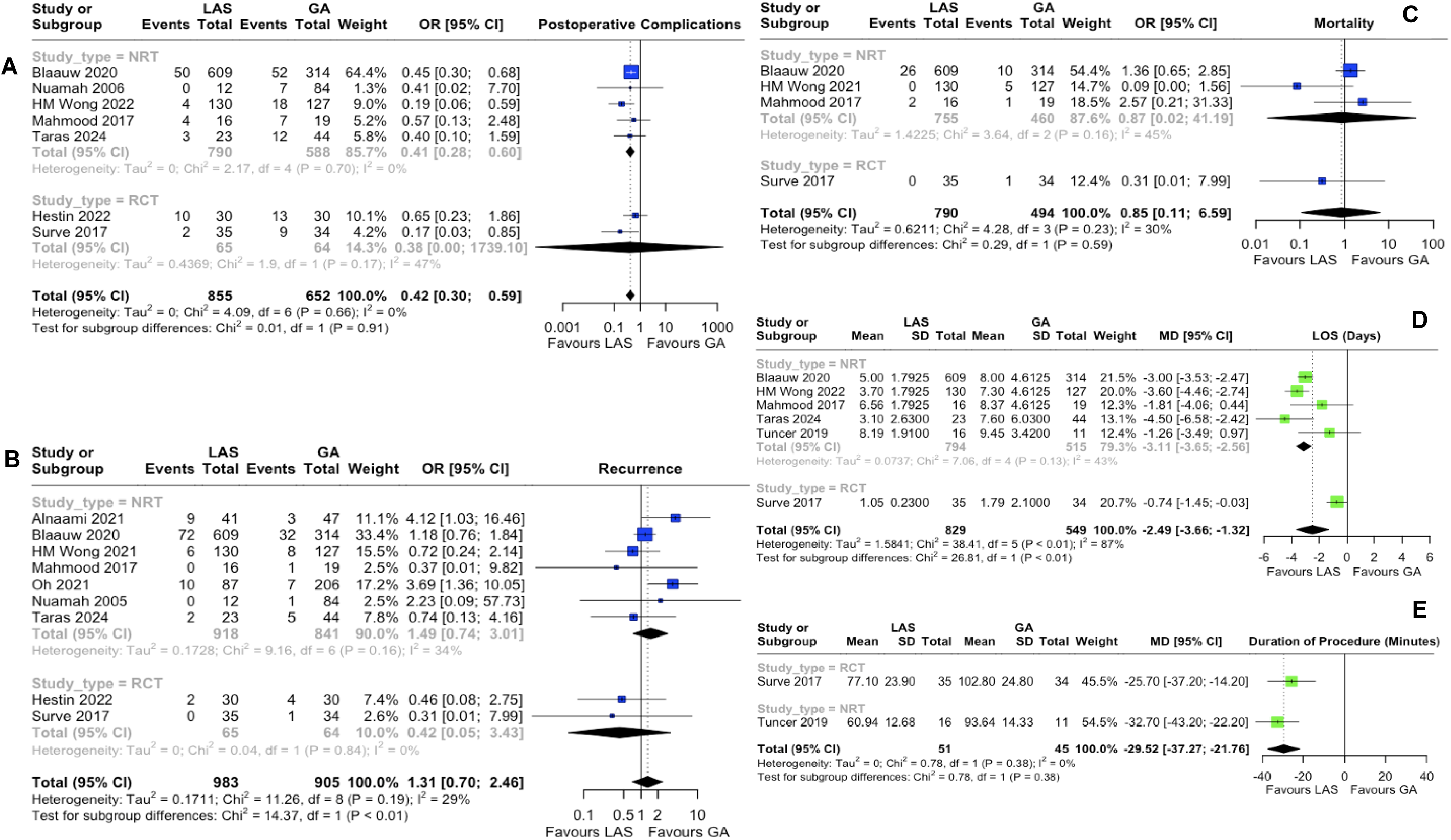
Forest plots for the meta-analyses of postoperative complications (a), recurrence (b), mortality (c), length of hospital stay (LOS) (d) and duration of anaesthetic and surgery (e) after studies that were at moderate to high risk of bias were removed. The studies are grouped based on whether they are NRTs or RCTs. Outcomes (odds ratio; blue, mean difference; green) are displayed. The number of individuals in each analysis is included. Heterogeneity statistics are demonstrated

### Ongoing studies on LA v GA in cSDH

We identified 11 registered trials of relevance **(Table 1)**. These were all single centre in nature and explored either LAS versus general anaesthesia (*n = 3*) or variants of LA technique (*n = 8*). These included comparisons of the use of pre-operative melatonin, dexmedetomidine/ketamine versus dexmedetomidine/propofol, or adjunctive magnesium sulphate versus fentanyl alongside propofol for sedation. Many trials comparing LAS techniques featured dexmedetomidine as a comparator arm (n=6). Postoperative inflammation is being assessed in one study [41].

**Table 1:** Registered clinical trials examining local anaesthetic (LA) techniques in chronic subdural haematoma (cSDH) surgery. In outcomes – bold = ‘primary’, non-bold = secondary. *ASDH = Acute subdural haematoma, HR = Heart Rate, ICU = Intensive Care Unit, mcg = micrograms, PE = Pulmonary Embolism, UTI = Urinary Tract Infection, VAS = Visual Analogue score,*

| Year registered | Status | Setting | Study Design | Outcomes |
| --- | --- | --- | --- | --- |
| 2024 [30] | Pending | Single-centre<br>Mansoura,<br>Egypt | *Comparison:* general anaesthesia or awake surgery with dexmedetomidine sedation (1mcg/kg over 10 minutes followed by 0.2-0.7mcg/kg/hour) in cSDH patients undergoing craniotomy<br>*Sample:* Target 62<br>*Inclusion:* Age $\geq 18$ and $\leq 80$<br>*Exclusion:* previous neurosurgery, other significant neurological disorder, severe medical comorbidities, allergies to study drugs, emergency surgery | • **Postoperative inflammatory markers (CRP, IL-6, IL-8)**<br>• Stress hormones (cortisol, noradrenaline) pre- and postoperatively<br>• Postoperative pain using VAS<br>• Time to discharge from hospital<br>• Postoperative complications<br>• Time to first rescue analgesia<br>• Postoperative mean arterial pressure<br>• Postoperative heart rate |
| 2024 [31] | Completed<br>– no<br>results<br>listed | Single-centre<br>Berlin,<br>Germany | *Comparison:* local anaesthesia or general anaesthesia<br>*Sample:* Target 50<br>*Inclusions:* Age $\geq 18$, confirmed symptomatic cSDH, informed consent | • **Recruitment rate** (consent rate and surgery rate)<br>• **Complication rate** (discharge and 1 month)<br>• Postoperative delirium<br>• GCS at discharge and 1 month<br>• Markwalder cSDH scale at discharge and 1 month<br>• Barthel Index at discharge and 1 month |
|  |  |  | *Exclusions:* bilateral cSDH, preoperative delirium, allergy to local anaesthesia or adrenaline, psychiatric contraindication to awake surgery, >24 hours after admission, CSF shunt present | • Mortality at 1 month<br>• Length of stay<br>• Discharge location<br>• Health economic analysis<br>• Reasons for non-participation |
| **2023 [32]** | Recruiting | Single-centre<br>Beijing, China | *Comparison:* Scalp block + dexmedetomidine (2-4mcg/kg loading over 10 minutes then 0.5-1mcg/kg/hr to achieve RASS-3)<br>*Sample:* Target 190<br>*Inclusions:* Age $\geq 18$ and $\leq 80$; haematoma thickness/shift > 1cm<br>*Exclusions:* GCS <13, Sensory or motor aphasia, recurrent, severe comorbidity/other organ dysfunction, severe coagulopathy | • **Intra-operative movement**<br>• Neurological function (discharge and 6 months)<br>• Conversion to GA<br>• Intraoperative awareness<br>• Postoperative delirium<br>• Recurrence at 6 months |
| **2023 [33]** | Complete<br>– no<br>results<br>listed | Single centre<br>Cairo, Egypt | *Comparison:* Scalp block with bupivacaine and lidocaine or scalp block with magnesium sulfate, bupivacaine and lidocaine in patients undergoing cSDH evacuation under general anaesthesia | • **Time to first dose of postoperative analgesia**<br>• Perioperative heart rate<br>• Perioperative systolic, diastolic and mean blood pressure |
|  |  |  | *Sample:* Target 38<br>*Inclusions:* ASA II-III, GCS > 13, age ≥ 45<br>*Exclusions:* patients needing postoperative ventilation, allergy to local anaesthetics, failed block, > 2 doses of rescue analgesia within the first postoperative hour, local infection, advanced cardiac pathology | • Postoperative pain using VAS |
| 2022 [34] | Not Yet Recruiting | Number of centres not specified<br>Varanasi, India | *Comparison:* Dexmedetomidine (1mcg/kg followed by infusion at 0.2-0.7mcg/kg/hr) or ketofol 1mg/kg loading dose followed by infusion or ketofol 0.5mg/kg/hour infusion in those undergoing burr hole evacuation of cSDH under monitored anaesthetic care<br>*Sample:* Target 60<br>*Inclusions:* Age ≥ 25 and ≤ 60, ASA I – III<br>*Exclusions:* Local infection at site of scalp block, uncontrolled hypertension, HR < 50, SBP < 90mmHg, DBP < 60mmHg | • **Intraoperative haemodynamics**<br>• Surgeon satisfaction |
| 2020 [35] | Completed<br>– no<br>results<br>listed | Single centre,<br>Zagazig,<br>Egypt | *Comparison:* Dexmedetomidine-Ketamine versus Dexmedetomidine-Propofol titrated to achieve modified observer's assessment of alertness and sedation score (OAA?S) 3<br>*Inclusions:* Age 50-80, BMI 25-30, ASA II/III, burr-hole evacuation<br>*Exclusions:* Difficult airway (Mallampati III/IV), altered mental status, PTSD, baseline sedative/hypnotic medications, heart block, severe liver/respiratory/renal impairment | • **Onset of sedation**<br>• Airway obstruction on a three-point scale<br>• Intraoperative rescue analgesia (fentanyl)<br>• Hypotension (MAP fall of > 20%), bradycardia (HR fall of > 20%), bradypnea (respiratory rate < 10), hypoxia (SpO2 <95%)<br>• Time to eye-opening<br>• Neurological status using Markwalder's Neurological Grading scale<br>• Surgeon satisfaction (7 point LIKERT) |
| 2019 [36] | Not Yet<br>Recruiting | Number of<br>centres not<br>specified<br>Kerala, India | *Comparison:* Scalp block + dexmedetomidine (0.2-0.5mcg/kg/hr) or propofol (25-50mcg/kg/min)<br>*Sample:* Target 250<br>*Inclusions:* Age > 65, ASA II and III, cSDH for burr hole evacuation<br>*Exclusions:* GCS < 15, repeat surgery, weakness of upper dominant limb, history of CVA, Parkinson's disease, cognitive decline > 3 months, patient<br>refusal, Mini Mental State Evaluation < 20 or Clinical Dementia Rating > 3 | • **Cognitive dysfunction** (2 days, 3 months, 1 year post-op)<br>• Functional outcome (2 days, 3 months, 1 year post-op)<br>• Haemodynamic and ventilatory parameters |
| **2018 [37]** | Unknown | Single centre, Caen, France | *Comparison:* Xylocaine 1% for scalp block versus General Anaesthesia<br>*Sample:* Estimated 60<br>*Inclusions:* Age ≥ 18, uni- or bilateral cSDH, French speaker<br>*Exclusions:* Agitated/non-cooperative, other intracranial lesions, underlying neurological pathology, pregnant/lactating women | • **Postoperative hospitalisation**<br>• Modified Rankin Score at 3 months<br>• Surgical complications: ASDH, infection, hydrocephalus<br>• Medical complications: Pneumonia, UTI, phlebitis, PE, other<br>• Patient/surgeon satisfaction (Likert 5 point)<br>• Cognitive impairment (MINI COG) at 6 months<br>• Mortality at 6 months<br>• Numeric pain rating scale at 2 days incl. opioid consumption<br>• ICU admissions |
| **2018 [38]** | Completed<br>– no results listed | Single centre, Cairo, Egypt | *Comparison:* Pre-operative oral melatonin versus placebo in those undergoing cSDH drainage under LA<br>*Inclusions:* Age 50-65, ASA I – II, Unilateral, stable vital signs, GCS 14-15<br>*Exclusions:* GI symptoms, substance abuse, hepatic/renal insufficiency, antipsychotic/anticonvulsant/anti-parkinson medications; unable to tolerate propofol sedation | • **Propofol consumption (in milligrams)**<br>• Blood pressure<br>• Intraoperative patient movement<br>• Time to recover from anaesthesia<br>• Visual analogue score (VAS) pain<br>• Time to first rescue analgesic |
| 2018 [39] | Completed<br>– no<br>results<br>listed | Single centre,<br>Kasr El Aini,<br>Egypt | *Comparison:* Non-inferiority comparison of magnesium sulphate versus fentanyl in comparison with propofol/lidocaine. Ramsay sedation score (RSS) of 3, with BIS sedation score of 60-80<br>*Inclusions:* Age > 50, ASA I-II, GCS 14-15, Unilateral cSDH<br>*Exclusions:* Hypertension, Bradycardia, Ischaemic heart disease (in last 6 months), second/third degree heart block, Substance abuse, allergy, neuropsychiatric diseases, predicted difficult airway (Ganzouri score > 4) | • **Propofol consumption**<br>• Patient movements<br>• Intraoperative/postoperative haemodynamics<br>• VAS for pain<br>• Time to rescue analgesic<br>• Adverse events<br>• Surgeon satisfaction |
| 2014 [40] | Completed<br>– no<br>results<br>listed | Number of<br>centres not<br>specified<br>Nihon, Japan | *Comparison:* dexmedetomidine versus propofol sedation for cSDH surgery under local anaesthesia<br>*Sample:* Target 40<br>*Inclusions:* Age >20, < 80<br>*Exclusions:* liver or renal dysfunction, arrhythmia | • **Efficacy and side effects** (details not specified) |

Importantly, most studies had restrictive inclusion criteria, excluding patients with significant baseline comorbidities, extremes of age, predicted airway difficulty, or low pre-operative GCS **(Table 1**). Outcomes were principally related to adequacy of technique (such as intra-operative movements or need for adjunctive propofol). Three studies stated they would examine surgeon satisfaction [35, 37, 39]; only one reported the intention of measuring patient satisfaction.

### Relevant studies in other conditions

Our search of the Cochrane database identified 12 systematic review records, although 3 (covering surgery for endovascular aortic aneurysm repair, vitrectomy, and cervical dilatation surgery) identified no relevant trials. Full details of identified Cochrane reviews are available in **supplemental table 4.** Major results, grouped by surgical discipline are shown in **Table 2**

**Table 2:** Summary of Cochrane reviews examining locoregional anaesthesia and general anaesthesia.

| **Surgical discipline** | **Number of studies** | **Study comparators** | **Outcomes examined** | **Findings** |
| --- | --- | --- | --- | --- |
| Breast Cancer | 9 – 7 in meta-analysis | Paravertebral or other truncal anaesthetic vs general anaesthesia | Quality of recovery<br>Pain<br>Block complications<br>Disease-free survival<br>Chronic pain<br>Postoperative quality of life | Regional anaesthesia may reduce pain and postoperative nausea and vomiting |
| Carotid Endarterectomy | 16 – 14 in meta-analysis<br>Dominated by the GALA trial of 4389 participants | Any locoregional technique (e.g. superficial/deep cervical plexus blocks) | Stroke<br>Death<br>Myocardial infarction<br>Haemorrhage<br>Cranial nerve injuries<br>Blood pressure<br>Patient/surgeon satisfaction | No difference except for a reduction in arterial shunting with local anaesthetic techniques |
| Caesarean section | 22 | Neuraxial | Death<br>Wound/other infections<br>Blood loss/change in Hb/Transfusion<br>Neonatal outcomes<br>Intraoperative pain<br>Postoperative pain/PONV<br>Maternal satisfaction | Reduced blood loss with neuraxial anaesthesia<br>More women using GA said they would use the same technique again in subsequent LSCS |
| General Surgery | 9 Cochrane reviews, 40 studies | Neuraxial techniques for any surgical procedure either as a sole comparator or adjunct to general anaesthesia | Mortality<br>Pneumonia<br>Myocardial Infarction<br>'Serious Adverse Events' | Reduced 30 day mortality in those with neuraxial anaesthesia if intermediate/high cardiac risk.<br>Reduced risk of pneumonia |
| Incomplete miscarriage | 7 (although no high quality RCTs) | Paracervical local anaesthesia | Patient satisfaction<br>Pain<br>Mortality | Patient-centered outcomes NOT assessed despite being highly relevant |
| Hip Fracture | 31 – 28 in meta-analysis | Neuraxial<br>Peripheral nerve block (e.g. lumbar plexus block) | All cause mortality<br>Pneumonia<br>Myocardial Infarction<br>CVA<br>Acute Confusion<br>DVT<br>Length of hospital stay<br>Analgesia | Reduced DVT in the absence of intensive anticoagulation with regional anaesthesia. Unilateral spinal may reduce intraoperative hypotension |
| Lower-limb revascularisation | 4 | Neuraxial | Mortality<br>CVA<br>Myocardial Infarction<br>Nerve dysfunction<br>Amputation<br>Hospital stay<br>Postoperative cognitive dysfunction<br>Pneumonia<br>Recovery room complications<br>Participant satisfaction<br>Pain | Reduced rates of pneumonia with neuraxial anaesthesia |
| Paediatric herniorraphy | 5 | Regional anaesthesia (epidural, spinal, caudal) | Postoperative apnoeas/desaturation<br>Neurodevelopment<br>Failure rates<br>Operator satisfaction<br>Hospital/NICU/PICU stay | Reduced postoperative apnoeas if patients receiving sedation were excluded. Risk of failure with regional techniques |
| Radial Fracture | 3 | Nerve block<br>Haematoma block<br>IV Regional<br>anaesthesia | Failure rates<br>Anatomic alignment<br>Clinical outcomes<br>Resource use | Insufficient<br>evidence |

The 9 Cochrane reviews with data covered a variety of surgical procedures including hip fracture, carotid endarterectomy, general surgery, lower-limb revascularisation, gynaecological procedures, caesarean section, and breast cancer surgery. In total 163 randomised trials were included across these Cochrane reviews. Trials involved a multitude of comparisons dependent on the type of surgery including locoregional anaesthetic techniques and neuraxial blocks (e.g. spinal and epidural).

### Survey findings

We received 215 responses (n = 125 for anaesthesia, n = 89 for surgery). Denominator data is difficult to obtain but for surgery, and using published figures of approximately 1000 members, this would amount to a response rate of approximately 9% for surgeons. General anaesthesia was the most common technique reported by 88% of surgeons and 66% of anaesthetists. Local anaesthesia was standard practice for only 6% of anaesthetic respondents. Both groups indicated that they believed LA could offer benefits in terms of length of stay and postoperative complications.

Analysis of free-text comments (see **supplemental methods**) indicated that the main perceived advantage of LA over GA was that it permits surgery in those with significant cardiorespiratory comorbidity. However, this comes at the risk of intraoperative movement which could be worse in those with pre-operative agitation. Participants also appeared to indicate that LA may be less suitable for more invasive or prolonged surgical procedures. Importantly, concerns about a longer recovery profile with LAS were raised as well as a potentially harmful effect on patient experience.

Both surgeons and anaesthetists appeared to feel that comorbidities, frailty and increasing age favoured the use of LA, while airway concerns and agitated behaviour strongly favoured general anaesthesia (**figure 3**). In general, views of anaesthetists and surgeons aligned, except on night-time operating (anaesthetists trend towards favouring GA in such a situation) and unit policies/antithrombotic use (surgeons trend towards favouring GA) [11].

**Figure 2:**
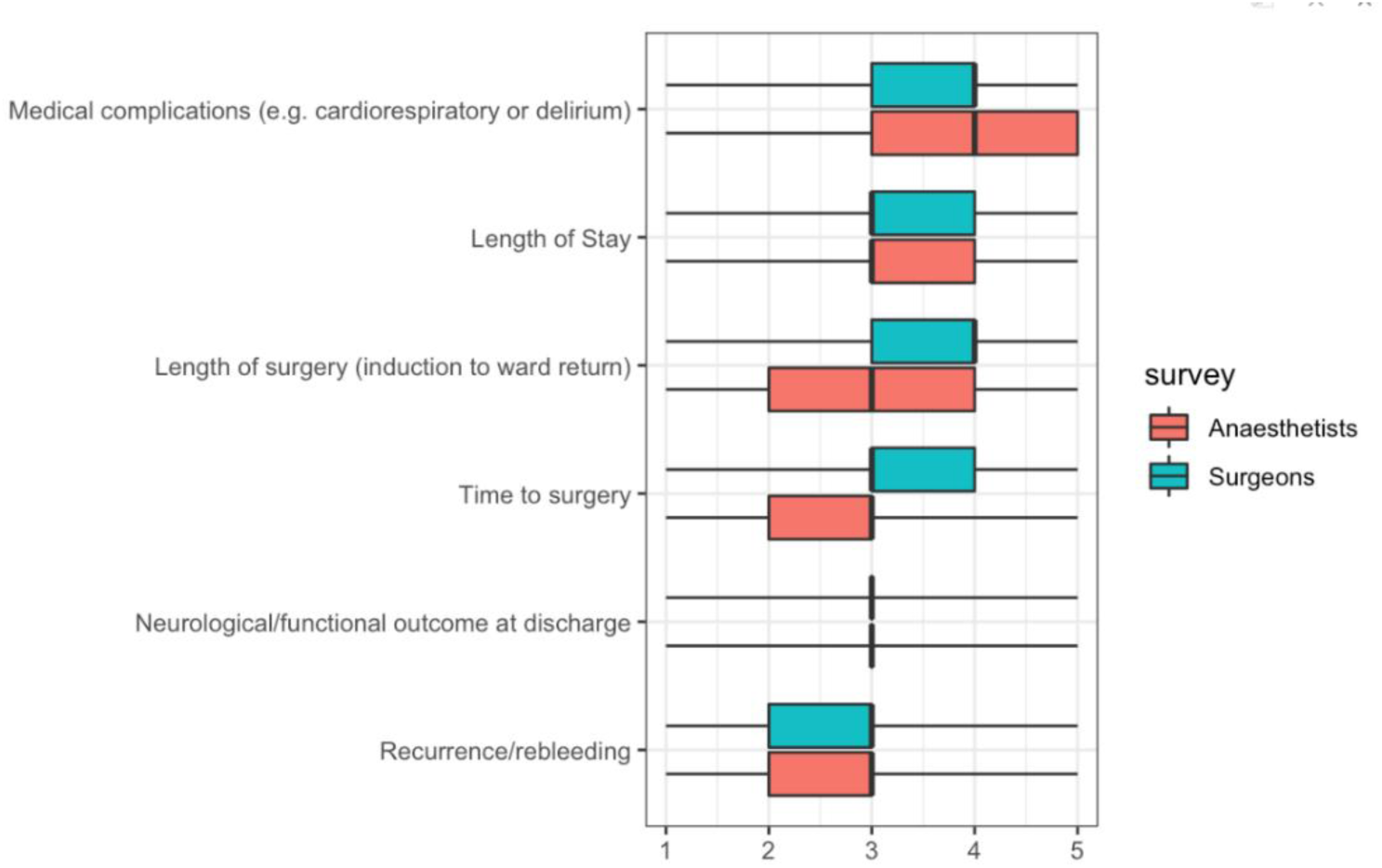
Perspectives of anaesthetists and surgeons on relation of postoperative outcomes to mode of anaesthesia (*1 = strongly favours general anaesthesia, 3 = equipoise, 5 = strongly favours local anaesthesia).* Box plots indicate mean and interquartile range.

**Figure 3:**
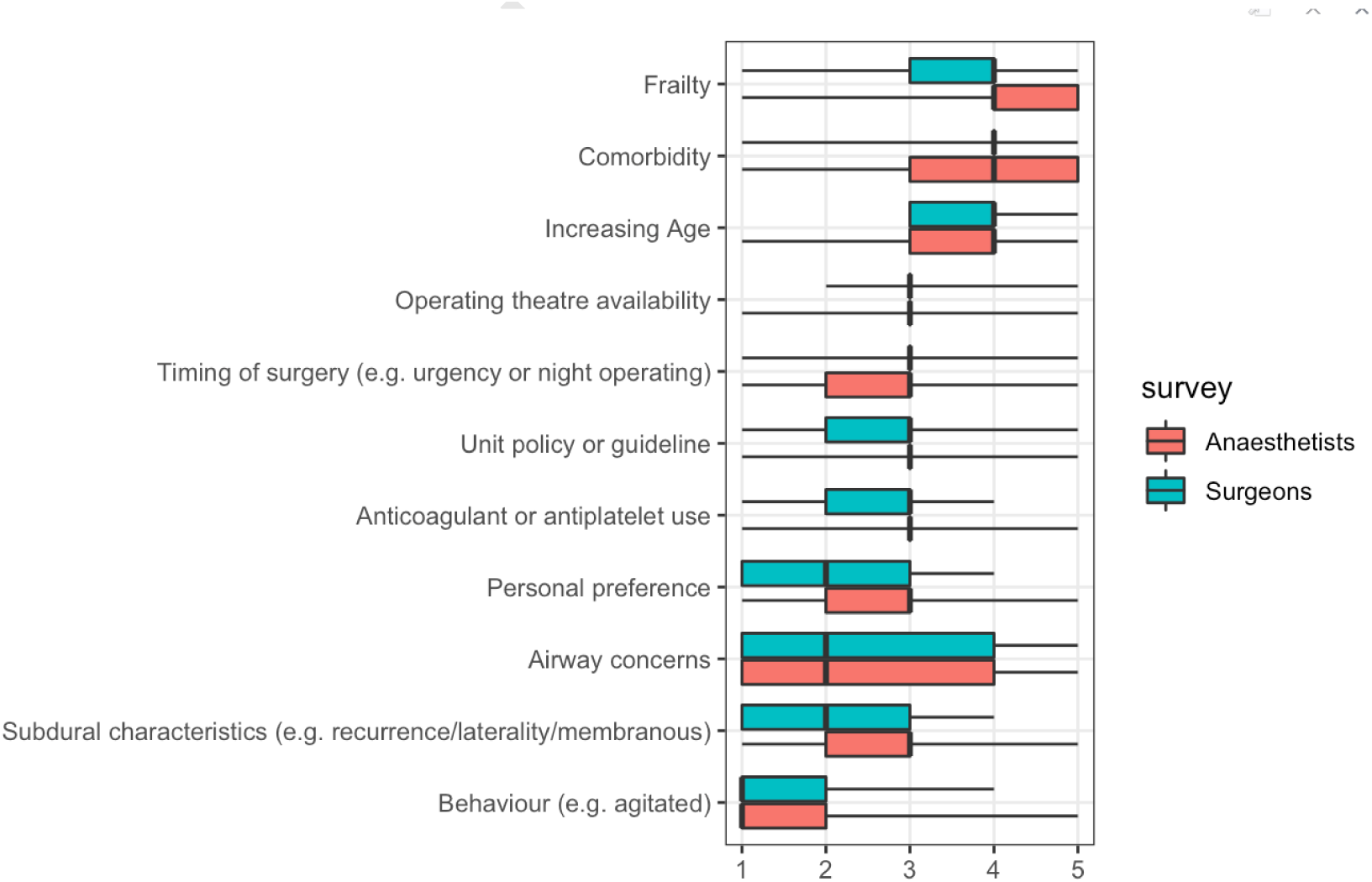
Perspectives of anaesthetists and surgeons on factors influencing choice of anaesthetic technique based on patient and operative characteristics. *(1 = strongly favours general anaesthesia, 3 = equipoise, 5 = strongly favours local anaesthesia)*.

We also thematically grouped free-text responses to questions. These themes were mapped as to whether they appeared to favour general or local anaesthesia (**figure 4**). These findings support the perception that LAS may bring benefits in terms of safety profile for high-risk individuals but that respondents indicate a lack of familiarity with LAS techniques, sub-optimal operative conditions, and tolerance.

**Figure 4:**
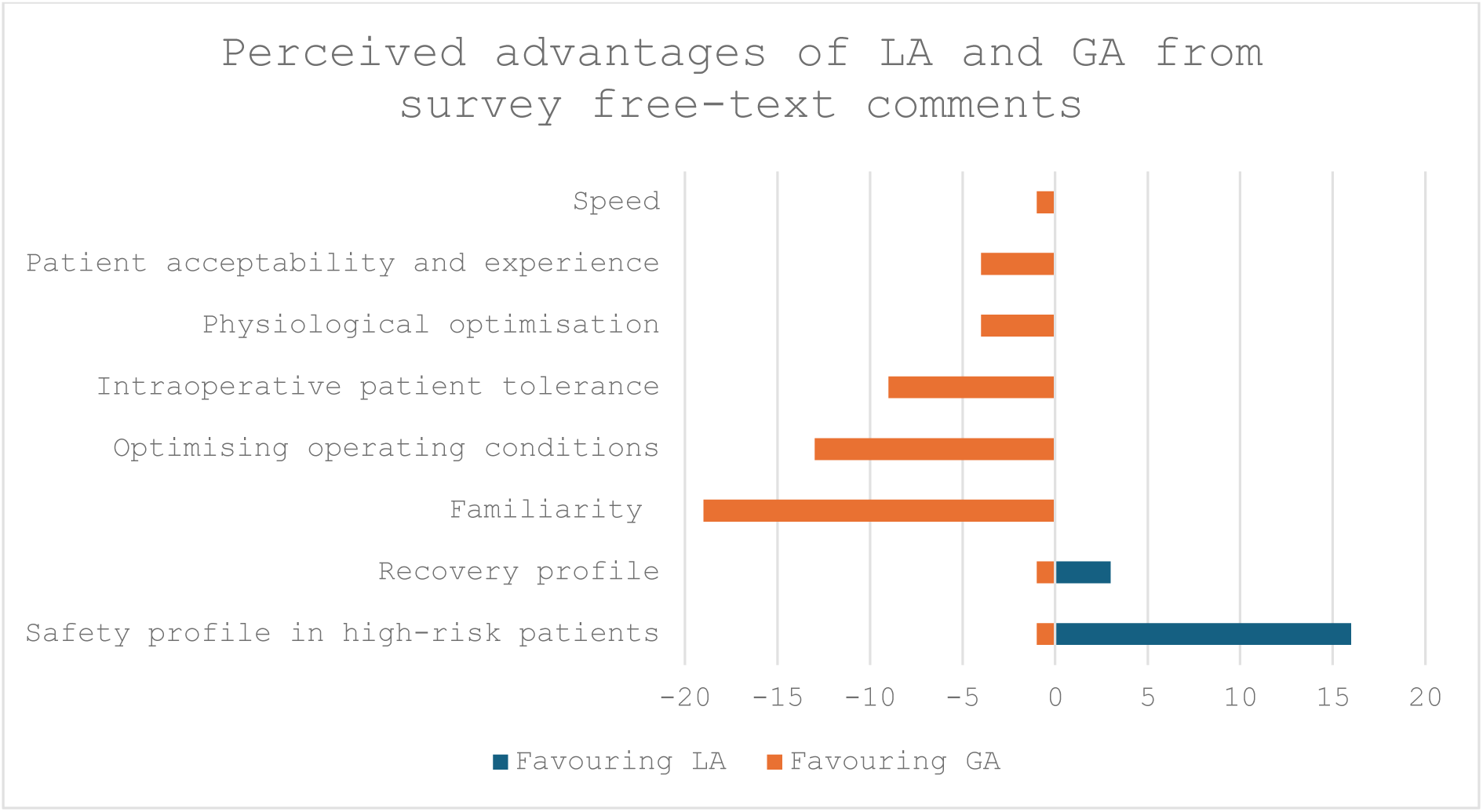
Major themes of anaesthetic and neurosurgical survey respondents on advantages of local anaesthesia (LA) versus general anaesthesia (GA). Numbers refer to number of free-text comments received mapping to each theme.

Sedation with propofol alone was the commonest adjunctive LA technique (23%). 53% of respondents chose intravenous maintenance for GA. 61% of respondents routinely used additional monitoring (26% arterial-line, 47% processed EEG). 61% of anaesthetic and 59% surgical respondents agreed that this question could benefit from further investigation with a randomised controlled trial. Importantly for any future trial a significant minority (∼20%) of anaesthetic respondents indicated that they felt ‘not confident’ or ‘probably not confident’ in delivering LAS for cSDH surgery.

## What could a definitive trial look like?

### Why do we need a randomised trial?

There remains a paucity of high-quality evidence to inform choice of anaesthetic technique; the vast majority of published studies are observational, retrospective, and/or single-centre. Clinicians are also uncertain, with variation in practice and viewpoints, despite competency in both techniques.

That said, meta-analysis suggests LAS could hold benefits, in length of stay and post-operative complications. Although the length of stay finding was consistent across studies, it is notable for post-operative complications that in a subgroup analysis of the two (albeit small) RCTs there was no difference. This raises a question as to whether the apparent protective effect of LAS may simply reflect confounding. The survey data and analysis of registered trials would be consistent with this, suggesting LAS is favoured in those with better baseline neurology or smaller collections. Together this reinforces the need for a randomised comparison in a heterogenous patient group. In support of this, the majority of both anaesthetists and surgeons felt the choice anaesthetic modality needed answering in a definitive RCT.

### What outcome should be used?

Evidence based medicine strives to improve patient outcomes and informative research must therefore ensure outcomes are patient-centred. This has given rise to the inclusion of patient reported outcomes measures. The challenge of applying this philosophy in perioperative research is well documented [42]. A scoping review of patient relevant outcome sets from multiple disciplines emphasises that complications, adverse events, and pain feature commonly [43]. The use of surrogates, whilst often objective and statistically powerful, can be difficult to interpret. Composite end-points (such as ‘pooled complications’) may falsely assume equal importance of each component, which could hide opposite (but equally important) effects on individual components [42].

In our meta-analysis, and other systematic reviews, the consistent signal in favour of LAS was a reduction in LOS. The biological basis for this is unclear. Some of this reduction could be driven by the modest (but statistically significant) reduction in procedural duration demonstrated in meta-analysis of approximately 30 minutes (**figure 1**). This may also interact with better scheduling, if, for instance, use of LAS helped reduce waiting times for theatre as well as post-operative complications.

LOS has not been a prominent outcome measure in cSDH research, with most trials favouring functional outcomes such as the modified Rankin scale or radiological outcomes such as ‘recurrence’. However, LOS is still a relevant outcome. From a patient perspective, LOS is associated with deconditioning and greater post-operative complications. Work in other fields suggests patients view a short hospital stay as their second most important From a service perspective, reduced LOS has health-economic and productivity benefits. This is of increasing significance, given the strained health service [44] and projected increased demand [2].

One of the challenges for interpreting LOS in cSDH care, is that most patients’ care is split across secondary and tertiary hospitals. Length of stay can also underrepresent readmissions, for example with recurrence, and might be influenced by organisational challenges such as inter-service transfer.

An alternative approach, not examined in any of the identified cSDH literature, may be to evaluate ‘days-alive-and-at-home’ at either 30 or 90 days. This metric, validated in hip fracture, is viewed as patient-centric by simultaneously encompassing acute and subacute hospital stay, the impact of complications, mortality, and disability [42]. Theoretically it could be calculated from linkage of trial to routinely collected (such as hospital episode statistic) data, reducing follow-up costs.

Recently, the GAP trial (Gabapentin versus Placebo as an adjunct to multimodal pain regiments) has used LOS (measured from the start of surgery to discharge) with secondary outcomes capturing clinical and patient quality of life metrics [45]. Their justification was that LOS served as an integrative measure of the overall potential of gabapentin to improve perioperative recovery through its (potential) effect on opioid reduction and thus complications [45]. A similar causal pathway could be postulated here, with impact on complications and/or recurrence being represented in a shorter LOS and higher number of days-alive-and-at home. Health related quality of life data is crucial to provide health economic assessment of this question. Patients with cSDH have reduced long-term survival, indicating the importance of a shorter LOS.

### Potential as a component of a platform trial

As highlighted through recent clinical practice guidelines [15], a significant number of questions exist in the care of cSDH that are amenable to well-conducted randomised trials. These include studies of medical interventions [46] as well as interventional radiological techniques such as middle meningeal artery embolization (MMAE) [18, 47]. Platform trials are *disease* rather than intervention focused, offering the opportunity for adaptive randomisation as best-practice is updated and offering significant efficiency gains through the sharing of infrastructure [46].

Given these benefits, the projected rise in cSDH cases, and current pressures on neurosurgical capacity there are many potential benefits of embedding a trial of LA v GA within a broader platform trial of cSDH care. Given the number of emerging therapies in this field this is a logical step. This design would also permit the assessment of pathway level interventions, especially relevant given the publication of new national guidance [15]. A platform design, alongside the use of routine hospital episode data [48] to capture longer term outcomes or embedding recruitment into data-collection for perioperative quality improvement should be considered, to maximise research efficiency and patient benefit.

### What is the patient experience of locoregional versus general anaesthesia?

No studies in cSDH formally examined patient satisfaction of surgery conducted under local as opposed to general anaesthesia, although one study identified on a clinical trials registry did report on planning to examine this. Failure rates of LAS of up-to 8% were reported in studies identified in our systematic search of cSDH **(Table 3)**.

Patient satisfaction of LAS for cSDH evacuation was evaluated in one study using measurements of intraoperative and postoperative pain, satisfaction, hospital anxiety and depression (HAD) scores, and Impact of Event Scale – Revised (IESR) [21]. 28 of 50 patients reported intraoperative pain (mainly headache) but at 3-5 days postop and at 6 months, upwards of 75% of patients would agree to undergo the procedure again under LAS. At immediate follow-up a small number of patients (n=3) met criteria for suspected post-traumatic stress disorder (PTSD) but this was not replicated at 6 months, although a similar number of patients had significant changes in their anxiety and depression scores at this point. Overall though, with good intraoperative communication and reassurance, these results suggest the procedure is relatively well tolerated.

Data from other conditions supports that overall locoregional anaesthesia is well tolerated, including in hand-surgery [49]. A prospective survey of patients undergoing carotid endarterectomy [50] demonstrated that older age and lower baseline cognition (assessed using the mini mental state examination – MMSE) had lower rates of pre-operative anxiety and lower requirements for preoperative information. This may be of relevance to cSDH surgery under LAS but formal examination of patient perspectives on acceptability are crucial. Our search of the Cochrane database (**supplemental table 4)** demonstrated that both patient and surgeon satisfaction had been poorly examined across other similar trials in other disciplines.

### What could a trial intervention look like? Currently used LAS techniques

Details of LAS techniques used in the literature are reported in **Table 3**.

Four studies compared different LAS techniques [13, 53, 54]. Dexmedetomidine was assessed in all studies. In each study, an alternative agent was used as comparator. These included fentanyl and midazolam, propofol, and sufentanil. One study assessed the use of two different doses of dexmedetomidine (Group D1 dexmedetomidine infusion at 0.5 μg kg^-1^; Group D2 dexmedetomidine infusion at 1 μg kg^-1^) [55]. In this study, dexmedetomidine at an infusion of 1 μg kg^-1^ was deemed to be superior to dexmedetomidine at an infusion of 0.5 μg kg^-1^ or sufentanil when assessed using intraoperative patient movements, rescue interventions, postoperative recovery and patient/surgeon satisfaction. Dexmedetomidine was also deemed superior to propofol [56] and fentanyl with midazolam [57, 58] when using similar metrics.

Although this low-quality data suggests potential benefits of dexmedetomidine, the effect of LAS on LOS was seen with other agents and propofol was the commonest agent reported as used in our survey (*23%*). Given the wide variation in practice, the pragmatic design choice would be to leave technique to the individual anaesthetist. Although more reflective of real-world practice, high quality process measurements or clear definitions would need to be taken to ensure group separation – perhaps using processed EEG metrics and defining a general anaesthetic as one where advanced airway management is required (such as a supraglottic airway device or endotracheal tube).

### What are the challenges of conducting a trial of anaesthetic modality?

The 9 Cochrane reviews we identified (**supplemental table 4**) demonstrated that similar questions of optimal anaesthetic technique, and the alternatives to general anaesthesia, have been explored across a variety of surgical disciplines. The rationale for this is perhaps clearest in trials in paediatric practice where the explicit aim has been to attempt to limit perceived harms of general anaesthesia both in the short-term (apnoea) [59] and longer-term (developmental harm) [60]. Similar questions of neurological side effects have prompted trials examining the impact of regional (spinal) *versus* general anaesthesia in patients with hip fracture (likely most comparable population) and also demonstrated no harm with the use of GA [54, 61].

Multiple lessons of relevance to any trial of LA vs GA in cSDH can be seen in this other literature **(Table 3)**. Beyond methodological issues of randomisation quality and assessor blinding, heterogeneity of both anaesthetic and surgical technique, as well as a lack of equipoise in putting ‘high-risk’ patients forward for randomisation were apparent in the GALA study of GA *versus* LA in carotid endarterectomy [62]. This might be especially relevant to cSDH given the potential for shared comorbidities between the cohorts and operative setup limiting access to the head/airway. A possible solution would be to control for this in trial inclusion criteria, identifying which modalities the patient may be eligible for on pre-operative assessment, before randomisation.

Many trials of locoregional *versus* general anaesthesia have permitted the co-use of sedative techniques as supplements. Being as many sedative agents can either be used as primary or adjunctive agents in general anaesthesia this creates the potential for contamination between comparison groups. Indeed, the RAGA trial in hip fracture surgery specifically examined spinal with no sedation as a comparator to general anaesthesia for this reason [54]. Future trials could include comparisons between GA, LA without sedation, and LA with titrated sedation, to avoid this contamination. This could incorporate main outcomes plus failure and conversion rates.

In terms of impact on outcomes few consistent signals can be seen although trials in lower-limb revascularisation and major surgical procedures highlighted a reduction in post-operative pneumonia rates with the use of neuraxial anaesthesia. However, this was not seen uniformly across trials of neuraxial anaesthesia (see **Table 3**). Also, it is vital to consider whether these findings reflect reductions in primary or secondary outcomes. Importantly no significant effect on longer term outcomes such as mortality were observed.

Taken together these issues mean that the choice of comparator and trial design is crucially dependent on the perceived causal relationship between exposure (LA technique) and outcome. A clear tension exists between a pragmatic design (clearly favourable in terms of recruitment given the spread of techniques in use in our survey and the literature – **Table 4**) and the risk for heterogeneity in exposure or poor group separation.

**Table 4.** Local anaesthetic techniques explored in the literature identified from a systematic review.

| Study/Year | Local anaesthetic technique | Surgical technique | Other details | Failure rate (% convert to GA) |
| --- | --- | --- | --- | --- |
| Ashry et al, 2022 | Scalp block and midazolam (2-4mg/hr) or propofol (1-3mg/kg/hr) | 2 x Burr hole |  | Not stated |
| **Bishnoi et al 2016 [51]** | Dexmedetomidine (1µg/kg over 10 minutes followed by continuous infusion 0.03 to 0.07 µg/kg/h) vs IV fentanyl 0.5 µg/kg and midazolam 0.03 mg/kg followed by 0.5 to 1.16 µg/kg/h fentanyl + 0.03 - 0.07 mg/kg/h midazolam<br>+ Lidocaine with adrenaline to incision site | Burr hole | Randomised (n=52)<br>Dexmedetomidine arm had fewer patient movement, better surgical satisfaction, and comparable patient satisfaction | 0% |
| **Hestin et al, 2022** | Scalp block and remifentanyl (0.5 – 1.5ng/ml) | 2 different methods used; burr hole craniostomy and craniotomy |  | 7% |
| **HM Wong et al, 2022** | Scalp block and fentanyl (0.5-0.75mg/kg) | Burr hole |  | 0% |
| **Mahmood et al 2017** | 1-2mg Midazolam then 2% Lidocaine without adrenaline | Mini craniotomy |  | Not stated |
| **Raja et al 2020** | Dexmedetomidine 1ug loading followed by 0.3 µg/kg/min + fentanyl 1µg/kg vs fentanyl 1µg/kg + midazolam 0.03mg/kg | Burr hole | Randomised n = 56.<br>Lower HR and blood pressure with dexmedetomidine.<br>Higher | Not stated |

|  |  |  | PaO2 in recovery |  |
| --- | --- | --- | --- | --- |
| **Srivastava et al 2018 [52]** | dexmedetomidine 1 $\mu\text{g kg}^{-1}$ over 10 minutes as a loading dose, followed by 0.2–0.7 $\mu\text{g kg}^{-1} \text{ hr}^{-1}$. Vs propofol 1 mg $\text{kg}^{-1}$ over 10 minutes as a loading dose, followed by 1–3 mg $\text{kg}^{-1} \text{ hr}^{-1}$ alongside scalp block. | Burr hole | Randomised. $N = 62$<br>One patient developed airway obstruction, two required vasopressors. Improved surgeon satisfaction with dexmedetomidine | 0% |
| **Surve et al, 2017** | Local infiltration and dexmedetomidine (1mcg/kg loading over 10 minutes followed by 0.5mcg/kg/hr) and fentanyl bolus 1mcg/kg) pre skin infiltration | Burr hole |  | 8% |
| **Taras et al, 2024** | Local infiltration (2% Lidocaine) and diazepam (no dose details) | Burr hole |  | Not stated |
| **Tuncer et al** | Fentanyl/Propofol/Midazolam/Dexmedetomidine (no details) | Unilateral double burr hole |  |  |
| **Wang et al 2016** | Dexmedetomidine at 0.5 $\mu\text{g kg}^{-1}$ for 10 min vs | Burr hole | Randomised. $N = 215$ | 0% |
| | Dexmedetomidine at 1.0 $\mu\text{g}\cdot\text{kg}^{-1}$ for 10 min vs Sufentanil 0.3 $\mu\text{g}\cdot\text{kg}^{-1}$ for 10 min | | across three arms. Greater rates of rescue midazolam in low dose Dex. And Sufentanil groups. Better patient and surgeon satisfaction at high dose of Dex. | |
| **Zhuang et al, 2022** | Local infiltration | Burr hole |  | 0% |

## Conclusions

Definitive evidence for a specific anaesthetic modality in cSDH surgery is lacking. There is a suggestion that LAS may offer advantages in terms of reduced complications and LOS, although the primarily observational nature of evidence means the risk of confounding is high. Using up-to-date meta-analysis, a survey of relevant practitioners, and a review of emerging and published literature demonstrates the need for a definitive randomised trial in this field. Based on this data a pragmatic approach to anaesthetic technique and a focus on patient relevant, outcomes such as days alive and at home would seem prudent.

## Data Availability

All data produced in the present work are contained in the manuscript

## Abbreviations and Acronyms

CSDH: chronic subdural haematoma
LAS: local anaesthetic with or without sedation
GA: general anaesthetic
RCT: randomised-controlled trial
NRT: non-randomised trial
NOS: Newcastle Ottawa Scale
ICENI: Improving Care in Elderly Neurosurgery Initiative

