## Supplementary material for "Need for a definitive trial of local versus general anaesthesia in chronic subdural haematoma; lessons from a systematic review, survey, and scoping review of other surgical conditions": x

**Authors and affiliations:**

Daniel J. Stubbs[1], Conor S. Gillespie[2,3], Matthew L. Watson[1], Basil Nourallah[1], Caroline M Phillips[1], George Gathercole[1], Jamie Brannigan[2,3], Keng Siang Lee[4], Orla Mantle[2,3], Vian Omar[2,3], Adele Mazzoleni[2,3], Githmi Palahepitiya Gamage[5], Alvaro Yanez Touzet[6], Munashe Veremu[2,3], Youssef Chedid[2,3], William H Cook[2,3], Karanjit Loyal[1], Gideon Adegboyega[2,3], Oliver D Mowforth[2,3], Edward Goacher[7], Apoorva Singh[1], Jonathan P. Coles[1], Alexis Joannides[2,3], Angelos Kolias[2,3], Judith Dinsmore[8], Iain Moppett[9,10], Michael Nathanson[10], Sally R Wilson[11], Amit Deshmukh[1], Edoardo Viaroli[2,3], David K. Menon[1], Ellie Edlmann[12], Benjamin M. Davies[2,3], Peter J Hutchinson[2,3], Improving Care In Elderly Neurosurgery Initiative (ICENI) Working Group

[1]Department of Medicine, Perioperative, Acute, Critical, and Emergency Care (PACE) Section, University of Cambridge, Cambridge, UK

[2]Department of Neurosurgery, Addenbrooke's Hospital, Cambridge, UK

[3]Department of Clinical Neurosciences, University of Cambridge, Cambridge, UK

[4]Department of Neurosurgery, King's College Hospital, London, UK; Department of Basic and Clinical Neurosciences, Maurice Wohl Clinical Neuroscience Institute, Institute of Psychiatry, Psychology and Neuroscience (IoPPN), King's College London, London, UK

[5]Royal College of Surgeons of Ireland, Dublin, Republic of Ireland

[6]School of Medical Sciences, Faculty of Biology, Medicine and Health, University of Manchester, Manchester, UK

[7]Department of Neurosurgery, Hull Teaching Hospitals, Hull, United Kingdom

[8]Department of Neuroanaesthesia, St George's Hospital, London, UK

[9]Academic Unit of Injury, Inflammation and Repair, University of Nottingham, Nottingham, UK

[10]Consultant anaesthetist, Department of Anaesthesia, Nottingham University Hospitals NHS Trust, Nottingham, UK

[11]Department of Neuroanaesthesia, University College London Hospitals NHS Foundation Trust, London, UK

[12]Department of Neurosurgery, South West Neurosurgical Centre, Plymouth, UK

DRAFT UNDER REVIEW

### Supplemental Methods

**Systematic review and meta-analysis of cSDH**

A systematic literature search was performed of Medline and Excerpta Medica Database (Embase), from inception to 2nd January 2024. Search strategy for both databases can be found below. We reviewed the bibliographies of included articles to identify additional studies. Papers were limited to English language. The PRISMA checklist can be found in supplementary materials. The study was registered with PROSPERO (registration number: CRD42022374873). Articles addressing this review question were identified in parallel with studies addressing other research priorities [1] to support new guideline development [2]. Full details of article screening and risk of bias assessment are available in supplementary methods.

We pragmatically defined LAS as any anaesthetic technique involving the use of local anaesthetic including direct infiltration (field block) or scalp block. Patients could receive sedative medications but could not require advanced airway management (defined as need for an endotracheal tube or supraglottic airway device). General anaesthesia was defined as a technique involving the use of an equivalent advanced airway device with anaesthesia maintained by intravenous or inhalational agents.

We included studies examining the impact of anaesthetic technique on; postoperative complications, recurrence, mortality, hospital stay, and duration of anaesthesia and surgery. Postoperative complications were extracted from the included articles and included medical or surgical events occurring post evacuation during the same hospital admission. Definitions of recurrence were pragmatic based on individual studies but broadly considered ipsilateral CSDH significant enough to require re-operation. Postoperative mortality was defined as death within 3 months of CSDH evacuation. Duration of anaesthesia and surgery was the total time spent in theatre to undergo anaesthetic and surgery for CSDH evacuation (or as defined by study authors).

All statistical analysis was carried out using R Version 4.0.2 [3], using the meta package. Odds ratio (OR) meta-analyses using the Mantel–Haenszel method were computed for postoperative complication rate, recurrence rate and mortality rate. Mean difference meta-analyses were computed for length of stay (LOS) and duration of anaesthesia and surgery. Sensitivity analyses were performed excluding studies deemed at risk of bias; including studies scoring a 'low risk of bias' on the Cochrane risk of bias tool-2 or scoring seven to nine on the Newcastle Ottawa Scale (NOS) [4, 5]. Subsequent sub-group analyses were then performed between RCTs and NRTs to identify any effect that may be only due to observational studies. Heterogeneity was measured by the $I^2$ test [6]. Funnel plots were used to assess publication bias, but were not generated if less than three studies were included [7].

DRAFT UNDER REVIEW

#### Search strategies

A comprehensive literature search was performed on 1st May 2022 of 2 databases: Medline, and Excerpta Medica Database (Embase), from inception. The exact search strategy for all databases and their results can be found below.

MEDLINE 28th April 2022

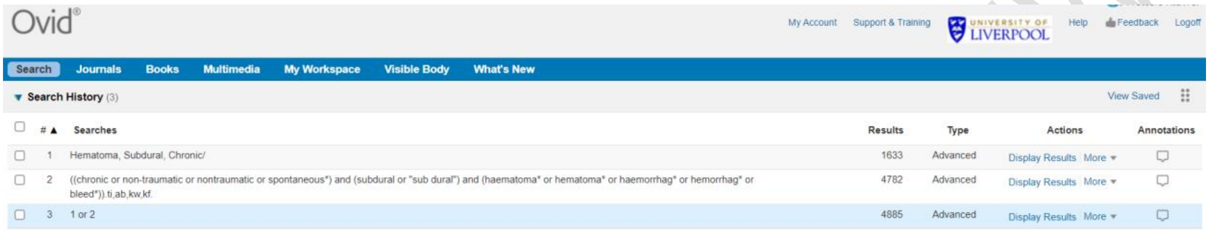
Ovid® My Account Support & Training UNIVERSITY OF LIVERPOOL Help Feedback Logoff

Search Journals Books Multimedia My Workspace Visible Body What's New

Search History (3) View Saved

| # | Searches | Results | Type | Actions | Annotations |
|---|---|---|---|---|---|
| 1 | Hematoma, Subdural, Chronic/ | 1633 | Advanced | Display Results More | |
| 2 | ((chronic or non-traumatic or nontraumatic or spontaneous*) and (subdural or "sub dural") and (haematoma* or hematoma* or haemorrhag* or hemorrhag* or bleed*)).ti,ab,kw,kf. | 4782 | Advanced | Display Results More | |
| 3 | 1 or 2 | 4885 | Advanced | Display Results More | |

Embase 28th April 2022

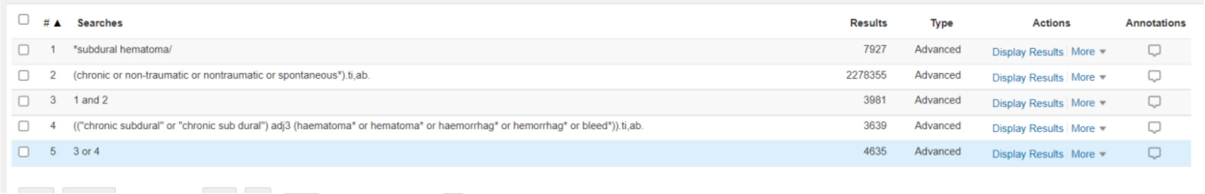
| # | Searches | Results | Type | Actions | Annotations |
|---|---|---|---|---|---|
| 1 | *subdural hematoma/ | 7927 | Advanced | Display Results More | |
| 2 | (chronic or non-traumatic or nontraumatic or spontaneous*).ti,ab. | 2278355 | Advanced | Display Results More | |
| 3 | 1 and 2 | 3981 | Advanced | Display Results More | |
| 4 | (("chronic subdural" or "chronic sub dural") adj3 (haematoma* or hematoma* or haemorrhag* or hemorrhag* or bleed*)).ti,ab. | 3639 | Advanced | Display Results More | |
| 5 | 3 or 4 | 4635 | Advanced | Display Results More | |

We reviewed the reference lists and bibliographies of included articles to attempt to identify additional studies. Papers were limited to English Language due to the difficulty in translation.

DRAFT UNDER REVIEW

Study screening and selection

All identified citations were transferred to the online platform Rayyan [8], a repository to facilitate removal of duplicate records and independent screening of potential records. Duplicates were then removed. This article superset was used to inform all systematic reviews for guideline development [1]. Initially, each of these articles were screened by two blinded, independent reviewers. Articles were excluded based on inclusion and exclusion criteria outlined in Supplementary File 3. Articles matching inclusion criteria were then categorised dependent on their title, journal, and abstracts, into pre-defined categories corresponding to specific systematic review questions. These categories included:

1. Anticoagulant
2. Decision Making
3. Communication
4. Anaesthesia and Surgical Scheduling
5. Transfer and Pathway
6. Perioperative Care
7. Palliative Care
8. Postop and recovery
9. Natural history
10. Surgical technique
11. MMA Embolisation

We excluded studies that were conference abstracts, case reports, and mixed populations where it was not possible to delineate CSDH-specific results (e.g., a dataset containing both acute SDH and chronic SDH data). These abstracts were then assessed again using the population, intervention, comparison, outcome, and study design (PICOS) criteria defined in Supplementary Table 1 by two independent, blinded reviewers (MW, CG). These criteria were selected to identify studies pertinent to our primary aim. Any other articles that did not meet these criteria, but contained relevant information surrounding anaesthetic methodology for CSDH evacuation that could be used to address our secondary aim were separately identified.

**Supplementary Table 1. PICOS inclusion criteria with definitions for the primary aim.** PICO question derived from the output of a cross-disciplinary working group.

DRAFT UNDER REVIEW

| | |
|---|---|
| **Review Question** | In adult patients undergoing surgical evacuation of a chronic subdural haematoma, does the use of local anaesthesia with or without sedation improve patient and system outcomes when compared to the use of general anaesthesia? |
| **Population** | Adult patients undergoing surgical evacuation of a chronic subdural haematoma |
| **Intervention** | The use of local anaesthetic with or without sedation for the operative procedure |
| **Comparator** | The use of general anaesthetic for the operative procedure |
| **Outcomes** | 1. Postoperative complication rate<br>2. Recurrence rate of subdural haematoma<br>3. Mortality rate postoperatively<br>4. Length of hospital stay<br>5. Duration of anaesthesia and surgery |
| **Study design** | Meta-analysis of defined outcomes using primary data from included studies |

If any disagreements occurred at either stage of screening, an attempt was made to resolve this between two review authors, and if discussion failed to lead to consensus, senior authors (BMD, DJS) were consulted for clarification. If any data was not present or available in the articles identified, corresponding authors were contacted via email to request the data.

<u>Data extraction and synthesis</u>

Data extraction was conducted independently and in duplicate by two authors using a standardised pre-piloted data collection proforma. Supplementary Files 4 and 5 are data collection proformas populated with relevant data for included studies.

**Supplementary Results**

Our initial search identified a superset of 6,049 primary research studies into CSDH 304 of these studies were of relevance to reviews within our ‘Anaesthesia and Surgical Scheduling’ theme. Following inclusion criteria, 140 articles were included in this study. Following free-text screening, 19

DRAFT UNDER REVIEW

studies relating to primary and secondary aims were identified (**supplemental table 2)**. The PRISMA flowchart is shown

DRAFT UNDER REVIEW

**PRISMA Diagram: Supplementary Figure 1**

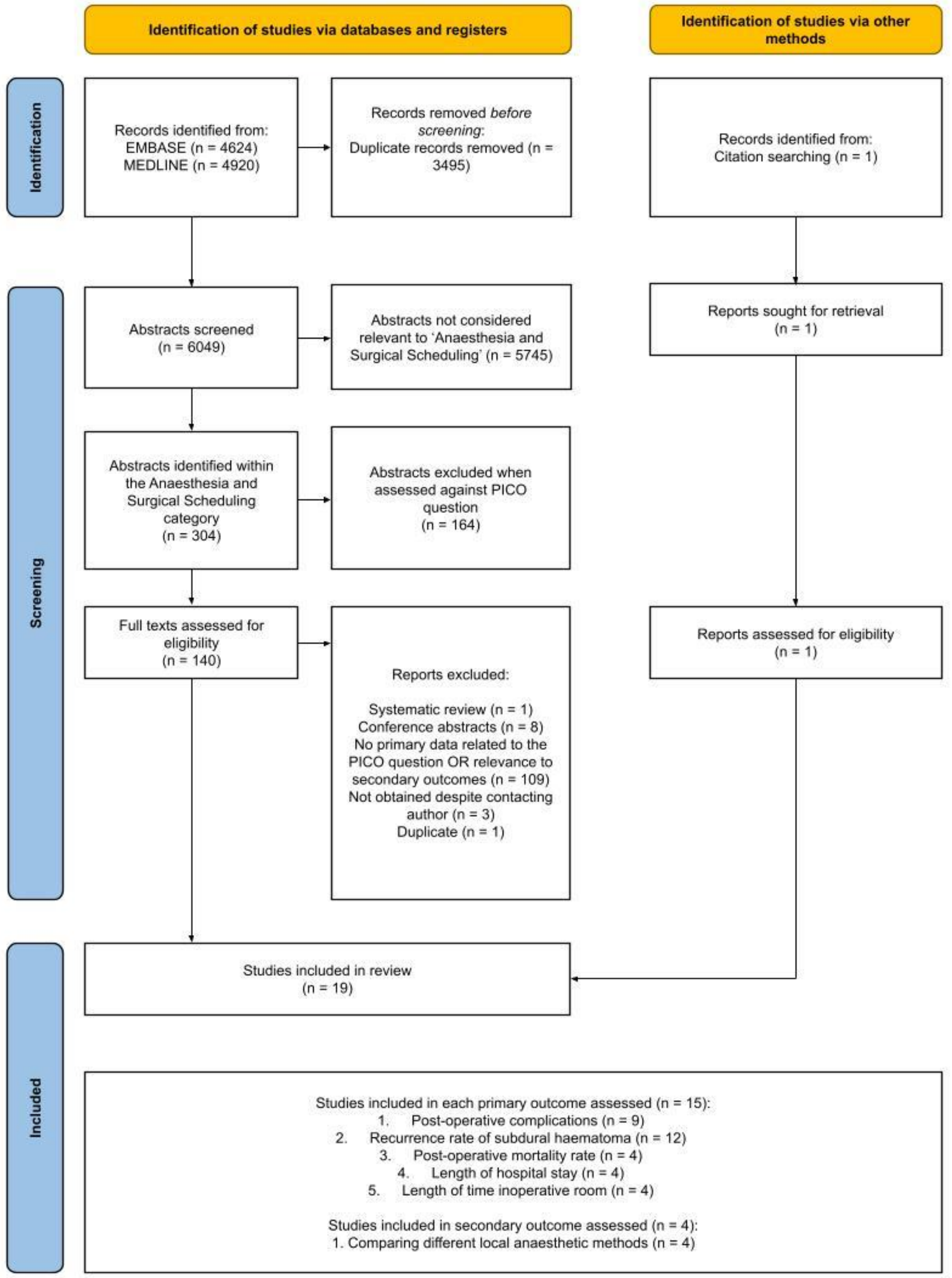

| Lead author of study | Type of publication | Year published | Risk of bias score | Country of the lead author | Number of cases | Number of LAS cases | Number of GA cases | Number of male cases | Mean age | Primary outcomes examined in study | | | | |
|---|---|---|---|---|---|---|---|---|---|---|---|---|---|---|
| | | | | | | | | | | Post-operative complications | Recurrence rate of subdural haematoma | Mortality rate post-operatively | Length of hospital stay | Length of time in theatre |
| Alnaami [9] | Retro and prospective cohort | 2021 | Low | Saudi Arabia | 88 | 41 | 47 | 71 | 64 | No data | Recorded and defined recurrence | No data | No data | No data |
| Ashry [10] | Retrospective cohort | 2022 | Medium | Egypt | 45 | 22 | 23 | 27 | 74 | Recorded postoperative complications | Do not define recurrence and no indication surgery was required | No data | Recorded mean length of hospital stay and standard deviation | Did not specify whether time recorded was including anaesthetic or not |
| Blaauw [11] | Retrospective cohort | 2020 | Low | Netherlands | 923 | 609 | 314 | 696 | 74 | Recorded postoperative complications | Recorded and defined recurrence | Recorded mortality data | Recorded mean length of hospital stay and standard deviation calculated | No data |
| Dakurah [12] | Retrospective cohort | 2006 | Low | Ghana | 96 | 12 | 84 | 90 | 47 | Recorded postoperative complications | Do not state which group had the recurrence | No data | No data | No data |

DRAFT UNDER REVIEW

| | | | | | | | | | | | | | | |
|---|---|---|---|---|---|---|---|---|---|---|---|---|---|---|
| Hestin [13] | Randomised control trial | 2022 | Low | France | 60 | 30 | 30 | 43 | 76 | Recorded postoperative complications | Recorded and defined recurrence | No data | Only reported median and IQR | Anaesthetic time not included |
| Hui Mei Wong [14] | Retrospective cohort | 2021 | Low | Malaysia | 257 | 130 | 127 | 201 | 68 | Recorded postoperative complications | Recorded and defined recurrence | Recorded mortality data | Recorded mean length of hospital stay and standard deviation calculated | No data |
| Mahmood [15] | Retrospective cohort | 2017 | Low | Pakistan | 35 | 16 | 19 | 25 | 69* | Recorded postoperative complications | Recorded and defined recurrence | Recorded mortality data | Recorded mean length of hospital stay and standard deviation calculated | Did not specify whether time recorded was including anaesthetic or not |
| Motiel-Langroudi [16] | Retrospective cohort | 2018 | Medium | America | 446 | 7 | 439 | 227 | 72 | No data | Data is based on re-operation (with other reasons such as acute SDH included) | No data | No data | No data |
| Oh [17] | Retrospective cohort | 2021 | Low | Korea | 293 | 87 | 206 | 208 | 75* | No data | Recorded and defined recurrence | No data | No data | No data |

DRAFT UNDER RE

| | | | | | | | | | | | | | | |
|---|---|---|---|---|---|---|---|---|---|---|---|---|---|---|
| Phang [18] | Retrospective cohort | 2015 | Low | United Kingdom | 275 | 70 | 205 | 187 | 79* | No data | Recorded and defined recurrence | No data | No data | No data |
| Surve [19] | Randomised control trial | 2016 | Low | India | 69 | 35 | 34 | 65 | 58 | Recorded postoperative complications | Recorded and defined recurrence | Recorded mortality data | Recorded mean length of hospital stay and standard deviation | Recorded anaesthetic + surgery time |
| Tuncer [20] | Retrospective cohort | 2019 | Low | Turkey | 27 | 16 | 11 | 15 | 75 | No data | No data | No data | Recorded mean length of hospital stay and standard deviation | Recorded anaesthetic + surgery time |
| Zhuang [21] | Retrospective cohort | 2022 | Low | China | 105 | 51 | 54 | 91 | 65 | Recorded postoperative complications | Recorded and defined recurrence | No data | Recorded time from operation to discharge | Recorded anaesthetic + surgery time |
| Certo [22] | Retrospective cohort | 2019 | Medium | Italy | 45 | 30 | 15 | 28 | 76 | No data | Recorded and defined recurrence | No data | Recorded time from operation to discharge | No data |
| Taras [23] | Retrospective cohort | 2024 | Low | Ukraine | 67 | 23 | 44 | 51 | 60 | Recorded postoperative complications | Recorded and defined recurrence | No data | Recorded mean length of hospital stay and | Did not specify whether time recorded |

DRAFT UNDER RE

| | | | | | | | | | | | | | | |
|---|---|---|---|---|---|---|---|---|---|---|---|---|---|---|
| | | | | | | | | | | | | | standard deviation | was including anaesthetic or not |

**Supplemental Table 2: Table describing characteristics of published studies comparing local versus general anaesthesia on pre-specified outcomes. The lead author of the study is provided with the reference. The year the study was published, and the country of the lead author is provided. The number of cases, number of GA/LAS cases, number of male patients and average age for each study is described. Whether a study was included in the assessment of a primary outcome is displayed.** *Median age; this was provided if the study did not provide a mean. LAS; local anaesthetic with or without sedation. GA: general anaesthetic. *Green box: study included in initial meta-analysis and data recorded. Yellow box: study contained data related to primary outcome but was not used any further for reason cited. Red box: no data relating to the specific outcome in question*

DRAFT UNDER REVIE

| Lead author of study | Postoperative complications examined in each study and their definition | | | | |
|---|---|---|---|---|---|
| | **Cardiac** | **Pulmonary** | **Neurological** | **Surgical** | **Other** |
| Ashry [10] | Hypertension<br>Acute coronary syndrome | - | Tension pneumocephalus<br>Seizures | Surgical site infection | Diabetic coma |
| Blaauw [11] | - | - | Delirium<br>Seizures | Rebleeding at wound site<br>Surgical site infection | Sepsis |
| Dakurah [12] | - | - | Tension pneumocephalus<br>Intracerebral haemorrhage<br>Cerebrospinal fluid leakage | - | - |
| Hestin [13] | Acute coronary syndrome | Dyspnoea | Delirium<br>Ischaemic stroke | Surgical site infection | Alcohol withdrawal<br>Urine infection |
| Hui Mei Wong [14] | Acute coronary syndrome | Pneumonia | Stroke<br>Intracerebral bleeds<br>Seizures | Surgical site infection | Thromboembolic events |
| Mahmood [15] | - | Pneumonia<br>Pleural effusion | Decreased GCS<br>Seizures | Rebleeding at wound site | Anaemia |

DRAFT UNDER REVIEW

| | | | | | |
|---|---|---|---|---|---|
| Surve [19] | Bradycardia | Dyspnoea | Delirium<br>Decreased GCS<br>Tension pneumocephalus | - | Stridor<br>Throat pain<br>Diarrhoea |
| Zhuang [21] | - | Pneumonia<br>Dyspnoea | Delirium<br>Decreased GCS<br>Seizures<br>Meningoencephalitis | Poor wound healing<br>Drain insertion into cerebrum | Vomiting<br>Sore throat |
| Taras [23] | - | - | Tension Pneumocephalus | - | - |

**Supplemental table 3 Table identifying the different postoperative complications described in the initial studies that analysed postoperative complications. The complications described have been categorised by major organ system into cardiac, pulmonary, neurological, surgical, and other.**

DRAFT UNDER REVIEW

| **Author/Study** | **Study design** | **Number of studies and patients** | **Outcomes and findings** | **Identified issues** |
|---|---|---|---|---|
| General versus loco-regional anesthesia for endovascular aortic aneurysm repair<br>Lee et al 2023<br>[24] | Types of studies: RCTs<br>Population: patients undergoing endovascular abdominal or thoracic aortic aneurysm repair<br>Intervention: loco-regional anaesthesia<br>Comparison: general anaesthesia<br>Primary outcomes: 30-day all-cause perioperative mortality, length of hospital stay, length of intensive care stay<br>Secondary outcomes: incidence of endoleaks, requirement for re-intervention, incidence of myocardial infarction, quality of life (e.g. SF-36), incidence of respiratory complications, pulmonary embolism and deep vein thrombosis, length of procedure | No eligible studies identified | None | Absence of well-designed RCTs on this topic |
| Local versus general anaesthesia for carotid endarterectomy<br>Rerkasem et al, 2021<br>[25] | Types of studies: RCTs<br>Population: people with symptomatic or asymptomatic carotid artery disease<br>Intervention: local anaesthesia (any locoregional technique)<br>Comparison: general anaesthesia | 16 studies (14 included in meta-analysis)<br>4839 participants | No significant difference in any stroke, death, ipsilateral stroke, stroke or death, myocardial infarction and other outcomes, except for a | Dominated by GALA trial (3526 of 4839 participants), which did not have a prescriptive study protocol leading to a **wide variation in anaesthetic technique** (any general anaesthetic vs any locoregional technique) and surgical practice.<br>In GALA, **higher-risk individuals**, who may have been felt to be better suited to a particular type of anaesthetic, **may not have** |

DRAFT UNDER REVIEW

| | | | | |
|---|---|---|---|---|
| | Primary outcome: stroke of any kind within 30 days, and during long-term follow-up.<br>Secondary outcomes: death, stroke or death, myocardial infarction, local haemorrhage, cranial nerve injuries, blood pressure, shunted arteries, hospital stay, participant and surgeon satisfaction, and feasibility of performing operation under local anaesthesia | | reduced rate of arterial shunting with local anaesthesia. | **been recruited**, as the overall stroke risks in the trial were very low<br>In some studies, **some randomised participants were removed from the analysis**, especially those who crossed over anaesthetic type<br>No studies able to blind participants or personnel to the mode of anaesthesia, introducing **high risk of performance bias**. 3 of 16 studies (including GALA) reported that outcomes were assessed by neurologists who were blinded to the type of anaesthesia used<br>**Variation in definition of some outcomes** (e.g. hypotension and hypertension) made these impossible to meta-analyse<br>Few trials assessed **surgeon or participant satisfaction** |
| Paravertebral anaesthesia with or without sedation versus general anaesthesia for women undergoing breast cancer surgery<br>Chhabra et al 2021<br>[26] | Types of studies: RCTs<br>Population: adult women undergoing breast cancer surgery<br>Intervention: paravertebral anaesthesia or other truncal regional anaesthesia, with or without sedation<br>Comparison: general anaesthesia<br>Primary outcomes: quality of recovery, postoperative pain at rest and on movement, mortality related to anaesthetic technique<br>Secondary outcomes: adverse events due to | 9 studies (7 included in meta-analysis)<br>614 participants | Paravertebral anaesthesia probably reduces 24-hour postoperative analgesic use, incidence of postoperative nausea and vomiting and pain at 2 hours, and may reduce pain at 24 hours at rest and pain on movement at 6 and 24 hours | **Variation in surgical procedure** between and within studies, ranging from lumpectomy to modified radical mastectomy and reconstructive breast surgery<br>**Variation in blocks performed** (single- or multiple-level, with or without catheters)<br>**Variation in depth of sedation**, **and variation in sedative agents**. in one study some patients in the paravertebral group received no sedation and some received conscious sedation.<br>No assessment of quality of recovery **using a validated questionnaire** |

DRAFT UNDER REVIEW

| | | | | |
|---|---|---|---|---|
| | paravertebral block, disease-free survival, chronic pain postoperatively, postoperative quality of life | | postoperatively compared to general anaesthesia. Reports of adverse events with paravertebral block were rare. | **No assessment of disease-free survival, mortality, chronic pain or quality of life**, important metrics in breast cancer surgery<br>**High (but unavoidable) risk of performance bias** due to inability to randomise patients. In 4 studies there was **uncertainty about blinding of outcome assessors**. |
| Local versus general anaesthesia for adults undergoing pars plana vitrectomy surgery<br>Lincina et al 2016<br>[27] | Types of studies: RCTs or cluster-RCTs<br>Population: adults undergoing pars plana vitrectomy<br>Intervention: general anaesthesia<br>Comparison: local anaesthesia (sub-tenon, peribulbar, retrobulbar, topical)<br>Primary outcome: intraoperative adverse effects which may have a lasting effect on final visual outcome<br>Secondary outcomes: postoperative adverse events, further application of local anaesthetic after start of surgery due to pain, patient satisfaction, degree of akinesia, surgical success | No eligible studies identified | None | Absence of well-designed RCTs on this topic |
| Anaesthesia for hip fracture surgery in adults<br>Guay et al 2016<br>[28] | Types of studies: RCTs<br>Population: patients 16 or older undergoing emergency hip fracture repair surgery<br>intervention: regional anaesthesia (neuraxial or peripheral nerve block such as lumbar plexus block), | 31 studies (28 included in meta-analysis) 3231 participants | No differences except for reduced deep venous thrombosis in the absence of potent thromboprophylaxis | **Suboptimal methodological rigour** in many trials, with respect to method used of randomization, concealment of allocation, **assessor blinding** and intention-to-treat analysis |

DRAFT UNDER REVIEW

| | | | | |
|---|---|---|---|---|
| | with or without sedation<br>Comparison: general anaesthesia<br>Primary outcomes: all-cause mortality at 30 days, pneumonia, myocardial infarction<br>Secondary outcomes: cerebrovascular accident, acute confusional state, deep vein thrombosis, return to home, congestive cardiac failure, acute kidney injury, pulmonary embolism, unsatisfactory surgical results, number of patients transfused, length of hospital stay, length of surgery, intraoperative hypotension, urinary retention, incomplete or unsatisfactory analgesia | | with regional anaesthesia, and reduced intraoperative hypotension if unilateral or incremental spinal anaesthesia is used | **Definition of outcomes and time points for assessment** varied widely between studies (e.g. acute confusional state, intraoperative hypotension)<br>**General anaesthetic techniques unrepresentative of contemporary practice** (such as halothane maintenance) may have impacted results, and a correlation was demonstrated between the effect size for mortality and date of publication, suggesting that a lower mortality rate associated with regional anaesthesia was more pronounced in older trials |
| Regional (spinal, epidural, caudal) versus general anaesthesia in preterm infants undergoing inguinal herniorrhaphy in early infancy<br>Jones et al 2015<br>[29] | Types of studies: RCTs or quasi-RCTs<br>Population: preterm infants born at less than 37 weeks' gestation undergoing inguinal hernia repair before 60 weeks' postmenstrual age<br>Intervention: any form of regional anaesthesia (epidural, spinal or caudal)<br>Comparison: general anaesthesia<br>Primary outcomes: apnoea, desaturations, use of postoperative respiratory support, neurodevelopmental state at two-year follow-up<br>Secondary outcomes: anaesthetic effectiveness with respect to operator satisfaction, anaesthetic failure, | 5 trials<br>152 participants | Significant reduction in postoperative apnoea in regional group once patients received preoperative sedation were excluded.<br>Increased risk of anaesthetic agent failure or anaesthetic placement failure in the regional group. | **Poor quality of many included studies** – small sample size, poor randomisation, uncertain allocation concealment, incomplete outcome assessment blinding, inadequacy of intention-to-treat analysis<br>**Use of sedation in the regional group impacted results,** increasing the incidence of postoperative apnoea– only on the exclusion of patients receiving preoperative sedation was a significant difference between the groups detected<br>Pre-specified definition of apnoea in many studies inconsistent with accepted clinical definition |

DRAFT UNDER REVIEW

| | | | | |
|---|---|---|---|---|
| | postoperative pain, duration of surgery, temperature on admission to recovery/NICU/PICU; duration of postoperative hospital stay, non-routine postoperative admission to PICU/NICU | | | |
| Neuraxial blockade for the prevention of postoperative mortality and major morbidity: an overview of Cochrane systematic reviews<br>Guay et al 2014<br>[30] | Types of studies: previous Cochrane systematic reviews<br>Population: patients of any age undergoing surgery<br>Intervention: neuraxial anaesthesia, or neuraxial anaesthesia plus general anaesthesia<br>Comparison: general anaesthesia alone<br>Outcomes: death, chest infection, myocardial infarction, serious adverse events | 9 Cochrane reviews<br>40 studies – half compared general versus neuraxial anaesthesia, the other half general plus neuraxial anaesthesia versus general anaesthesia alone | Neuraxial anaesthesia as a sole anaesthetic technique reduced 30-day mortality rate in patients undergoing procedures of intermediate to high cardiac risk, as well as reducing risk of perioperative pneumonia. | **Reporting of side effects and complications** related to anaesthetic techniques was incomplete<br>**Blinding was usually not used and was considered not feasible or realistic**, though some authors tried to insert a "sham" epidural catheter subcutaneously |
| Neuraxial anaesthesia for lower-limb revascularisation<br>Barbosa et al 2013<br>[31] | Types of studies: RCTs<br>Population: adults aged 18 or older undergoing lower-limb revascularisation surgery<br>Intervention: neuraxial anaesthesia<br>Comparison: general anaesthesia | 4 studies<br>696 participants | No difference observed in mortality rate, myocardial infarction and lower-limb amputation. Pneumonia | **Insufficient number of participants** to detect a meaningful difference in mortality<br>**Variations in anaesthetic technique incompletely reported** (e.g. spinal or epidural drugs, and level of blocks) |

DRAFT UNDER REVIEW

| | | | | |
|---|---|---|---|---|
| | Primary outcomes: mortality, cerebral stroke, myocardial infarction, nerve dysfunction, rate of lower-limb amputation<br>Secondary outcomes: duration of hospital stay, postoperative cognitive dysfunction, postoperative wound infection, pneumonia, complications in anaesthetic recovery room, participant satisfaction, postoperative pain score, transfusion requirement, urinary retention, claudication distance, pain at rest | | was less common at neuraxial anaesthesia than general anaesthesia. Evidence was insufficient for other outcomes. | **Outdated anaesthetic technique** reported in older studies, limiting generalisability to modern practice<br>**Risk of selection bias** as most included studies did not sufficiently describe how random allocation and allocation concealment were achieved<br>**Most studies had unclear or inadequate blinding** of participants, study personnel and outcome assessors |
| Paracervical local anaesthesia for cervical dilatation and uterine intervention<br>Tangsiriwatthana et al 2013<br>[32] | Types of studies: randomised or controlled clinical studies<br>Population: women undergoing cervical dilatation and uterine intervention<br>Intervention: paracervical local anaesthesia<br>Comparison: no treatment, placebo, other methods of regional anaesthesia, systemic sedation and analgesia, general anaesthesia<br>Primary outcomes: | 26 studies<br>2790 participants | No studies found comparing general anaesthesia to paracervical local anaesthesia | Absence of well-designed RCTs on this topic |
| Anaesthesia for evacuation of incomplete miscarriage<br>Calvache et al 2012<br>[33] | Types of studies: RCTs or cluster-RCTs<br>Population: patients with a diagnosis of incomplete miscarriage undergoing surgical evacuation<br>Interventions: any anaesthetic technique given preoperatively or intraoperatively (general anaesthesia, sedation/analgesia, regional and | 7 studies<br>800 participants | No studies found comparing general anaesthesia to locoregional anaesthetic techniques | Absence of well-designed RCTs on this topic<br>In other studies, **patient-centred outcomes such as patient satisfaction and quality of life were not assessed**, despite being highly relevant to mode of anaesthesia decisions in this procedure |

DRAFT UNDER REVIEW

| | | | | |
|---|---|---|---|---|
| | paracervical local block)<br>Comparison: any other anaesthetic technique<br>Primary outcomes: patient satisfaction, pain during or after surgical evacuation, maternal mortality<br>Secondary outcome: adverse events | | | |
| Regional versus general anaesthesia for caesarean section<br>Afolabi et al 2012<br>[34] | Types of studies: RCTs or quasi-RCTs<br>Population: women having elective or emergency caesarean section for any indication<br>Intervention: regional anaesthesia, whether spinal, epidural or any combination of both<br>Comparison: general anaesthesia<br>Primary outcomes:<br>Maternal: death, postoperative wound or other infections, difference between pre- and postoperative haematocrit or haemoglobin, blood loss > 500ml, mean blood loss, amount of blood transfused, number receiving postoperative blood transfusion<br>Neonatal: death, mean umbilical arterial or venous pH, mean neurologic and adaptive score, mean Apgar at one and five minutes<br>Secondary outcomes:<br>Maternal: incidence of intraoperative pain, maternal | 29 studies met inclusion criteria but only 22 contributed data<br>1793 participants | No significant difference in major maternal or neonatal outcomes.<br>Neuraxial anaesthesia associated with significantly lower estimated blood loss and drop in haematocrit.<br>Significantly more women who had general anaesthesia stated that they would use the same technique again in a subsequent caesarean section. | Many of the included studies were **underpowered**<br>**Patient satisfaction was rarely assessed**, despite being an important and clinically relevant metric in caesarean section<br>Only one study analysed data in an **intention-to-treat** manner<br>Many studies **did not report method of randomisation or allocation concealment** |

DRAFT UNDER REVIEW

| | | | | |
|---|---|---|---|---|
| | satisfaction, need for postoperative analgesia, incidence of postoperative nausea and vomiting, time to request postoperative analgesia, adverse events.<br>Neonatal: time to sustained respiration, need for oxygen or intubation, Apgar score of 4, 6 or 8 or less at 1 and 5 minutes, mean Apgar score at 1 and 10 minutes | | | |
| Anaesthesia for treating distal radial fracture in adults<br>Handoll et al 2002<br>[35] | Types of studies: RCTs or quasi-RCTs<br>Population: patients who had completed skeletal growth receiving conservative or surgical treatment for a distal radius fracture<br>Interventions: difference types of techniques of anaesthesia (nerve block, haematoma block, intravenous regional anaesthesia, sedation, general anaesthesia)<br>Outcomes: failed/inadequate anaesthesia, anatomical restoration, adverse effects attributable to anaesthetic technique, clinical outcomes, functional outcomes, resource use | Of these, 3 trials involved general anaesthesia (versus haematoma block, sedation and haematoma block plus sedation)<br>At least 1200 participants | Insufficient robust evidence to establish relative effectiveness of different methods of anaesthesia in the treatment of distal radius fracture | **Inadequate reporting of trial methodology** in many studies, such as failure to ensure or confirm the concealment of allocation in all but 2 trials<br>Some trials **did not report highly relevant long-term outcomes** such as inadequate fracture reduction and functional outcomes<br>Studies involving general anaesthesia and sedation **did not report quantitative data on recovery time**<br>**Some studies exhibited confounders** such as imbalances between groups in fracture type and expertise of involved health professionals |

**Supplemental table 4: Cochrane reviews examining studies of locoregional versus general anaesthesia in other surgical disciplines.**

DRAFT UNDER REVIEW


DRAFT UNDER REVIEW


DRAFT UNDER REVIEW
